## Supplementary information for "Insulin sensitivity in mesolimbic pathways predicts and improves with weight loss in older dieters"

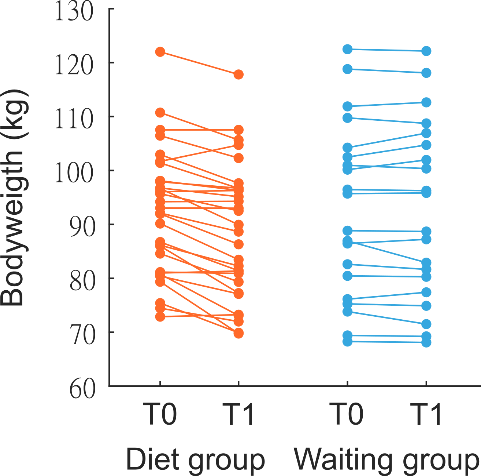

**Supplementary Figure 1. Individual weight-change in dieters and controls.** Weight-change in kg after three month dietary intervention / waiting phase.

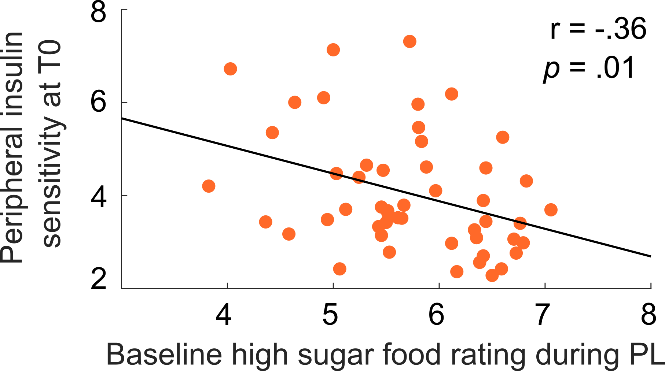

**Supplementary Figure 2. Correlation between sweet food liking and peripheral insulin sensitivity**. Lower insulin sensitivity as measured via the CPR-IR score was related to higher sugar liking. No correlation with insulin sensitivity was found for low sugar liking (P > .19).

| **Parametric liking score** | **Set 1** | **Set 2** | **Set 3** | **Set 4** | **P** |
| --- | --- | --- | --- | --- | --- |
| **All items** | 2.67 (0.05) | 2.73 (0.05) | 2.72 (0.05) | 2.77 (0.04) | NS |
| **Food items** | 2.86 (0.07) | 2.90 (0.07) | 2.98 (0.06) | 2.96 (0.06) | NS |
| **Non-food items** | 2.48 (0.05) | 2.56 (0.06) | 2.47 (0.06) | 2.59 (0.05) | NS |

**Supplementary Table 1. Validation of stimuli sets.** In a validation study, an independent sample of 16 participants rated the preference of items on a scale from 1 (~ “I do not like this at all”) to 4 (~ “I like this very much”). One-way ANOVAs showed that the four sets did not differ in regard to the parametric liking score across items and for food and non-food items separately. Values indicate means with s.e.m. in parentheses.

| **DG** | **T0** | | **P** | **T1** | | **P** | **P** |
| --- | --- | --- | --- | --- | --- | --- | --- |
|  | **PL** | **IN** | **(session)** | **PL** | **IN** | **(session)** | **(session x time)** |
| Fasting time (h) | 12.3 (0.3) | 12.2 (0.3) | NS | 12.4 (0.3) | 12.3 (0.3) | NS | NS |
| Hunger rating | 2.2 (0.4) | 2.3 (0.4) | NS | 2.9 (0.5) | 2.8 (0.5) | NS | NS |

| **WG** | **T0** | | **P** | **T1** | | **P** | **P** |
| --- | --- | --- | --- | --- | --- | --- | --- |
|  | **PL** | **IN** | **(session)** | **PL** | **IN** | **(session)** | **(session x time)** |
| Fasting time (h) | 12.4 (0.3) | 12.8 (0.4) | NS | 12.5 (0.3) | 12.6 (0.4) | NS | NS |
| Hunger rating | 2.5 (0.6) | 2.2 (0.5) | NS | 2.8 (0.5) | 3.1 (0.7) | NS | NS |

**Supplementary Table 2. Fasting duration before each study day and hunger ratings before each MRI scan.** Neither did the fasting times between the last food intake and the beginning of the study day differ between sessions (PL/IN, T0/T1) or groups, nor was there a group x session effect. Before entering the scanner, participants rated their current feelings of hunger on a scale from 0 (“not hungry at all”) to 10 (“extremely hungry”). Values did not differ between sessions or groups and there was no group x session interaction. Values indicate means with s.e.m. in parentheses. DG: diet group, WG: waiting group, PL: placebo, IN = insulin, T0: baseline, T1: follow-up. NS not significant.

| **DG_T0** | **PL** | | **P** | **IN** | | **P** | **P** |
| --- | --- | --- | --- | --- | --- | --- | --- |
|  | **Pre** | **Post** |  | **Pre** | **Post** |  | **(interaction)** |
| Insulin (pmol/L) | 78.8 (4.0) | 62.8 (4.6) | .003 | 80.6 (4.5) | 72.9 (6.0) | .16 | NS |
| Glucose (mmol/L) | 5.5 (.1) | 5.6 (.1) | NS | 5.4 (.1) | 5.4 (.1) | NS | NS |
| **DG_T1** |  |  |  |  |  |  |  |
| Insulin (pmol/L) | 70.3 (4.4) | 57.7 (3.8) | .001 | 72.9 (4.9) | 62.6 (4.9) | .036 | NS |
| Glucose (mmol/L) | 5.4 (.1) | 5.4 (.1) | NS | 5.5 (.1) | 5.3 (.1) | .02 | .049 |

| **WG_T0** | **PL** | | | **P** | **IN** | | **P** | **P** |
| --- | --- | --- | --- | --- | --- | --- | --- | --- |
|  | **Pre** | | **Post** |  | **Pre** | **Post** |  | **(interaction)** |
| Insulin (pmol/L) | 97. 9 (8.6) | | 70.2 (8.1) | .001 | 88.6 (7.4) | 70.2 (6.5) | .001 | NS |
| Glucose (mmol/L) | 5.7 (.1) | | 5.7 (.1) | NS | 5.7 (.1) | 5.6 (.1) | NS | NS |
| **WG_T1** |  | |  |  |  |  |  |  |
| Insulin (pmol/L) | 97.3 (7.9) | 65.3 (6.0) | | .001 | 103.0 (8.9) | 80.5 (7.8) | .001 | NS |
| Glucose (mmol/L) | 5.8 (.1) | | 5.7 (.1) | NS | 5.9 (.1) | 5.7 (.1) | .03 | NS |

**Supplementary Table 3. Pre-post blood values at baseline and follow-up.** Blood samples were sampled after arrival and after completion of the scanning sessions (see Figure 1b). There was a significant insulin level x session interaction across participants at T0 (F(1,49) = 4.1; P = .047, rmANOVA) driven by a stronger insulin decrease in the placebo session. This effect was not significant within single groups, in interaction with groups, nor were there any significant session effects at T1 (all P > .30). Values indicate means with s.e.m. in parentheses. DG: diet group, WG: waiting group, PL: placebo, IN = insulin, T0: baseline, T1: follow-up. NS not significant.

| **Dependent variable** | **Model (R² adjusted)** | **Predictor variable (standardized β –coefficients)** | | |
| --- | --- | --- | --- | --- |
|  |  | *Insulin sensitivity (CPR-IR)* | *Insulin effects on VTA signal* | *BMI* |
| Model 1 |  |  |  |  |
| *BMI change (%)* | .39*** | .36** | .54*** |  |
| Model 2 |  |  |  |  |
| *BMI change (%)* | .37** | .39* | .51** | .08 |

**Supplementary Table 4. Regression models for the prediction of dietary success.**

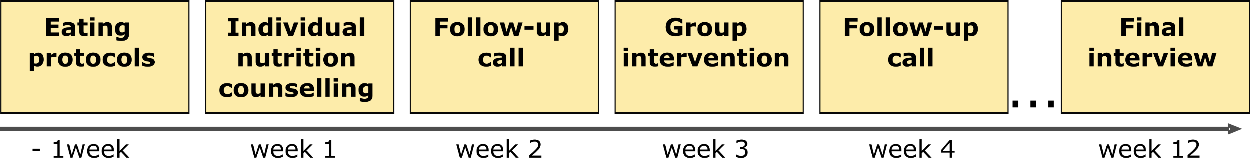

**Supplementary figure 3. Roadmap dietary intervention.**
